# Pathogens detected using a syndromic molecular diagnostic platform in patients hospitalized with severe respiratory illness in South Africa in 2017

**DOI:** 10.1101/2021.11.10.21266173

**Authors:** Malefu Moleleki, Mignon du Plessis, Kedibone Ndlangisa, Cayla Reddy, Anne von Gottberg, Orienka Hellferscee, Omphe Mekgoe, Meredith McMorrow, Sibongile Walaza, Cheryl Cohen, Stefano Tempia, Ebrahim Variava, Nicole Wolter

## Abstract

**Background:** Pneumonia continues to be a leading cause of death globally; however, in >50% of cases, an etiological agent is not identified. We describe the use of a multi-pathogen platform, TaqMan array card (TAC) real-time PCR, for the detection of pathogens in patients hospitalized with severe respiratory illness (SRI).

**Methods:** We conducted prospective hospital-based surveillance for SRI among patients at two sentinel sites in South Africa between January and December 2017. Patients were included in this study if a blood specimen and at least one respiratory specimen (naso- and oro-pharyngeal (NP/OP) swabs and/or sputum) were available for testing. We tested respiratory specimens for 21 respiratory pathogens and blood samples for nine bacteria using TAC. Pathogen detection was compared by age group and HIV status using the chi-squared test.

**Results:** During 2017, 956 patients were enrolled in SRI surveillance, and of these, 637 (67%) patients were included in this study (637 blood, 487 NP/OP and 411 sputum specimens tested). At least one pathogen was detected in 83% (527/637) of patients. Common pathogens detected included *H. influenzae* (225/637; 35%), *S. pneumoniae* (224/637; 35%), rhinovirus (144/637; 23%), *S. aureus* (129/637; 20%), *K. pneumoniae* (85/637; 13%), *M. tuberculosis* (75/637; 12%), and respiratory syncytial virus (57/637; 9%). Multiple pathogens (≥2) were co-detected in 57% (364/637) of patients.

**Conclusion:** While use of a multi-pathogen platform was useful in the detection of a pathogen in the majority of the patients, pathogen co-detections were common and would need clinical assessment for usefulness in individual-level treatment and management decisions.

## Background

Worldwide, pneumonia remains the leading cause of morbidity and mortality in children aged <5 years, accounting for 16% of all deaths between 2000 and 2016 (1). This burden is geographically disproportionate: low and middle-income countries accounted for 89% of the estimated 336 million pneumonia cases in all ages in 2016, and 62% of the estimated 2 million pneumonia deaths occurred in sub-Saharan Africa and South Asia (2). HIV infection is an important risk factor for pneumonia, specifically in South Africa where HIV prevalence among individuals hospitalized with pneumonia can be up to 74% in patients aged ≥5 years and 90% in adults aged 25-44 years (3, 4). Rapid identification of the etiological agent would allow for pathogen-directed treatment, in line with current antibiotic stewardship practices (5).

Conventional microbiological testing for pneumonia-causing pathogens involves use of culture, microscopy, antigen-antibody tests and pathogen-specific polymerase chain reaction (PCR) (6) assays (7-9). Through the severe respiratory illness (SRI) surveillance programme in South Africa, the etiology of community-acquired pneumonia for selected pathogens, namely influenza A and B viruses, parainfluenzavirus (PIV) 1–3, respiratory syncytial virus (RSV), enterovirus, human metapneumovirus (hMPV), adenovirus, human rhinovirus and *Streptococcus pneumoniae* using individual real-time PCR assays has been described (3, 4). However, a pathogen remained undetected in more than 50% of patients (3, 4). Multi-pathogen real-time PCR platforms may improve the detection of pathogens when compared to conventional tests (10, 11), and potentially highlight important roles of other pneumonia-causing pathogens in the era of the pneumococcal conjugate and *Haemophilus influenzae* type b vaccines (12, 13). In this study, we describe the use of the multi-pathogen platform, TaqMan array card (TAC) real-time PCR system, for the detection of pathogens in patients hospitalized with SRI.

## Materials and Methods

### Study population

Prospective hospital-based syndromic surveillance for SRI was initiated in 2009 in South Africa to describe the etiology of and risk factors for community-acquired pneumonia as previously described (14). Cases enrolled at two sentinel sites hospitals in the per-urban area of the North-West Province and a rural area of Mpumalanga Province in South Africa between January and December 2017 were included in the current study.

#### Case definition

A case of SRI was defined according to an adapted World Health Organization (WHO) case definition (14). Briefly, SRI was defined as any hospitalized individual, regardless of symptom duration, in age-defined categories as follows: any child aged 2 days to <3 months with diagnosis of suspected sepsis or physician diagnosed lower respiratory tract infection (LRTI) irrespective of signs and symptoms, any child ≥ 3 months to <5 years with physician-diagnosed acute LRTI including bronchiolitis, pneumonia, bronchitis and pleural effusion, and for older children aged ≥5 years and adults, as measured fever (>38°C) or reported fever and cough. All SRI cases with ≥10 days symptom duration were classified as chronic SRI. Surveillance officers administered a questionnaire with demographic information and obtained clinical information from medical records. In the event that routine HIV testing was not done, HIV testing was performed at National Institute for Communicable Diseases (NICD) in Johannesburg, South Africa as previously described (4).

### Specimen collection

Nasopharyngeal aspirates were collected from children aged <5 years and combined naso- / oro-pharyngeal (NP/OP) swabs from patients aged ≥5 years. Induced sputum specimens were collected from individuals aged ≥3 months. If induced sputum collection was contraindicated or was not advised by the attending clinician and a patient was able to expectorate, expectorated sputum was collected. Whole blood was collected from patients of all ages. Specimens were transported to the NICD for testing and thereafter stored at -80°C.

### Laboratory testing

#### Individual real-time PCR (IRTP) testing of SRI specimens

Total nucleic acid (15) was extracted from 200 µl each of blood, NP/OP and sputum using the Roche MagNA Pure 96 automated extractor (Roche Diagnostics, Mannheim, Germany) with the DNA and Viral Nucleic Acid Small Volume kit according to manufacturer’s instructions. In-house real-time PCR assays were carried out for the detection of *S. pneumoniae* in blood and *Bordetella* spp. (*B. pertussis, B. parapertussis* and *B. holmesii*) in NP/OP and sputum specimens (16, 17). A commercial Fast-Track Diagnostics FLU/HRSV real-time PCR kit (Fast-Track Diagnostics, Esch-sur-Alzette, Luxembourg) was used for the detection of influenza A viruses, influenza B viruses and respiratory syncytial virus in NP/OP specimens as previously described (18).

#### TAC testing

A total of 956 patients were enrolled in the SRI surveillance program, of which 637 were included in this study if a blood specimen and at least one respiratory specimen (naso- and oro-pharyngeal (NP/OP) swabs and/or sputum) were available for testing. From these patients, we tested any left-over specimens with sufficient volume: 200 µl for NP/OP and sputum specimens, and any volume for blood specimens (Fig 1).

**Fig 1:**
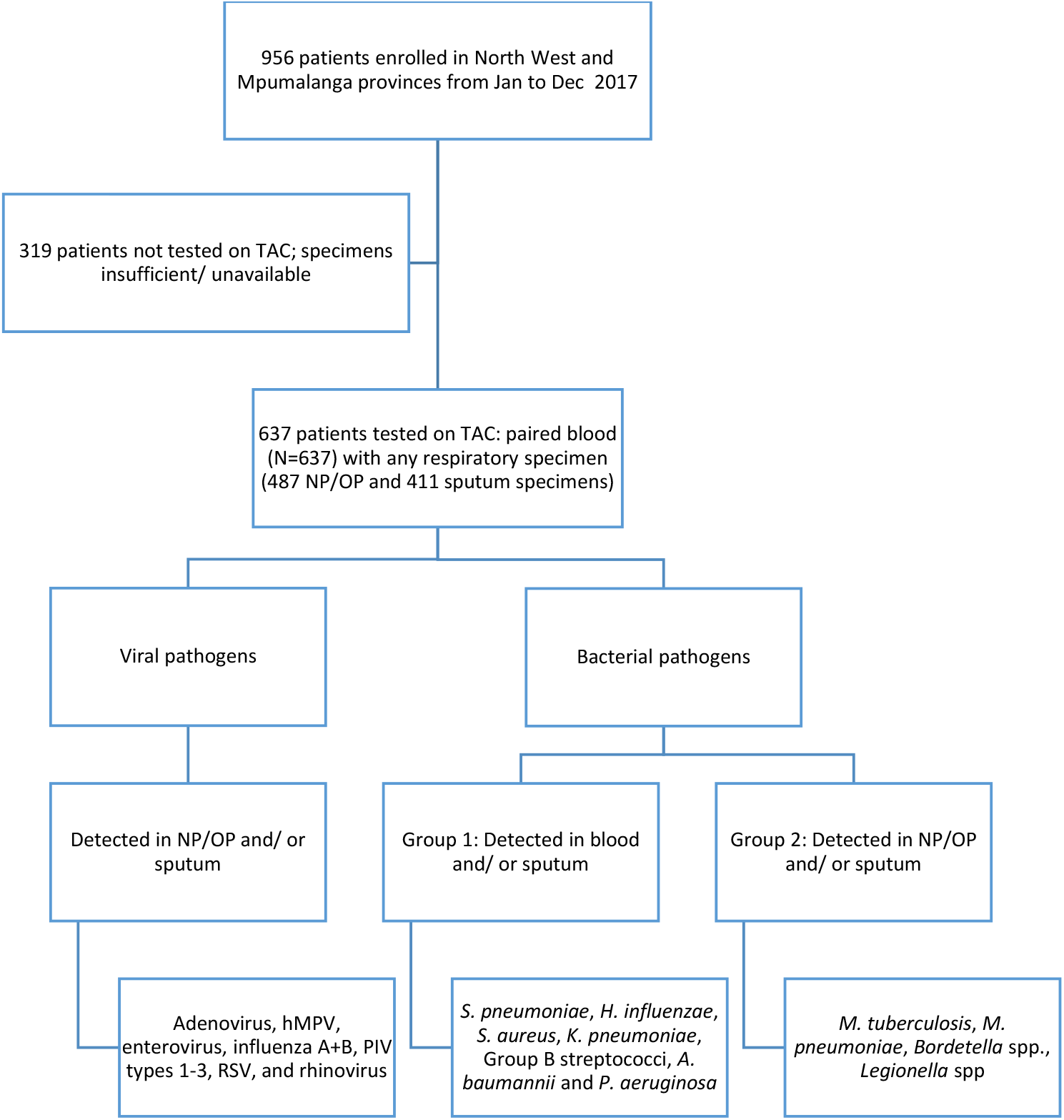
Algorithm for pathogen identification in patients hospitalized with severe respiratory illness tested on TAC who were enrolled at two sentinel sites in South Africa between Jan and Dec 2017. We excluded the following from further analysis: (A) cytomegalovirus detected in blood as detection of the pathogen DNA in blood cannot differentiate between latent and active disease. (B) *S. aureus, S. pneumoniae, H. influenzae, A. baumannii, K. pneumoniae, P. aeruginosa* and Group B streptococci if detected in NP/OP specimens as these are common colonizers of the upper respiratory tract and, (C) *E. coli/Shigella* spp. due to poor assay specificity. hMPV, human metapneumovirus. PIV, parainfluenza.

#### Nucleic acid extraction for TAC testing

An aliquot of 300 µl whole blood was incubated at 37°C for 60 min with an enzyme lysis solution (1.5 mg/mL lysostaphin, 2500 U/mL mutanolysin, and 200 mg/mL lysozyme (Sigma-Aldrich, St. Louis, MO, USA) in TE buffer prior to extraction (19). TNA was extracted using the MagNA Pure Compact instrument (Roche Diagnostics) with Total Nucleic Acid Isolation Kit I according to the manufacturer’s instructions. TNA was extracted from 200 µl of NP/OP and sputum specimens using the Roche MagNA Pure 96 automated extractor (Roche Diagnostics) with the DNA and Viral Nucleic Acid Small Volume kit according to manufacturer’s instructions.

#### TAC design

TaqMan Array Cards were adapted from previously published assays (11, 19, 20). Table S1 lists the organisms tested on the blood and respiratory TACs in this study. All targets were detected in two replicates on the respiratory TAC or four replicates on the blood TAC with the exception of *H. influenzae*, which was detected in two replicates on the blood TAC. Cards included two controls, namely -the internal positive control (IPC) for monitoring of the real-time PCR reaction and a human RNAseP gene control for monitoring of specimen quality.

#### TAC real-time PCR

Whole bloods were tested using the blood TAC while NP/OP specimens and sputum were tested using the respiratory TAC as previously described (19). Briefly, TAC assays were performed using 50 µl qScript XLT 1-step RT-qPCR Toughmix (Quantabio, Beverly, Massachusetts, USA) and 50 µl of TNA extract. TAC PCR was carried out on the Applied Biosystems ViiA7 and QuantStudio 7 Real Time PCR systems (Life Technologies, New York, USA) using the following cycling conditions: 45 °C for 10 min, 94 °C for 10 min, 45 cycles of 94 °C for 30 sec and 60 °C for 1 min. A no template control (NTC) and positive control consisting of combined RNA transcripts generated as previously described by Kodani et al. (2011) were included on each TAC (21). A positive result was recorded if amplification occurred in at least one of the duplicate or quadruplicate reactions with cycle threshold (Ct) <40.

#### Pathogen identification

The algorithm for pathogen identification is illustrated in Figure 1. Briefly; pathogen identification was defined as: (A) detected in NP/OP and/or sputum for *Mycobacterium tuberculosis, Mycoplasma pneumoniae, Bordetella* spp., *Legionella* spp., and all viruses, or (B) detected in blood and/or sputum for *Streptococcus pneumoniae, Haemophilus influenzae, Staphylococcus aureus, Klebsiella pneumoniae*, Group B streptococci (GBS), *Acinetobacter baumannii* and *Pseudomonas aeruginosa*. The following pathogens were excluded from analysis: cytomegalovirus (CMV) detected in blood as detection of the pathogen DNA in blood cannot differentiate between latent and active disease (22), *S. aureus, S. pneumoniae, H. influenzae, A. baumannii, K. pneumoniae, P. aeruginosa* and GBS if detected in NP/OP specimens as these are common colonizers of the upper respiratory tract (12, 23-25), and *E. coli/Shigella* spp. due to poor assay specificity (19, 20).

### Statistical analysis

Stata 14 (Stata Corporation, College Station, TX) was used for statistical analysis. McNemar’s χ2 test or Fischer’s exact test were used, where appropriate, to compare categorical variables (p-value <0.05 was considered statistically significant). The positivity proportion of *S. pneumoniae* in blood, *Bordetella* spp. in NP/OP and/or sputum specimens, and influenza A and B viruses, and RSV in NP/OP specimens were compared between TAC and IRTP assays. Concordance between the two tests was calculated using overall percent agreement (OPA). Agreement was further defined using Cohen’s *kappa* statistic (*κ*) which ranges between 0 and 1 in which *κ* = 0 indicates no agreement and *κ* = 1 is perfect agreement (26).

### Ethics statement

The national syndromic surveillance for pneumonia protocol (M140824) was approved by the Human Research Ethics Committee (HREC) of the University of the Witwatersrand as well as the local ethics committees of the respective sites. This surveillance was deemed non-research by the Centers for Disease Control and Prevention, Atlanta, USA (U.S. CDC) and did not need human subjects review by that institution. The protocol for the current study (M1902109) was approved by the HREC of the University of the Witwatersrand.

### Availability of data

All data generated or analysed during this study are included in this published article and its supplementary information files.

## Results

### Study population

From January to December 2017, 956 patients were enrolled in the surveillance study at the two sentinel sites and of these, 637 were selected for testing using TAC (Table 1). The median age of the patients tested on TAC was 33 years (interquartile range (IQR) 6-47 years), and 24% (154/637) were aged <5 years. SRI presentation was acute in 45% (286/637) of the patients, and 58% (369/637) were HIV-infected. The median duration of symptoms was 6 days (IQR 2-18 days). The case-fatality ratio was 7% (42/624) amongst the SRI patients with specimens tested using TAC.

**Table 1:** Demographic and clinical characteristics of severe respiratory illness (SRI) patients enrolled at two sentinel sites in South Africa, January – December 2017

| **Characteristic** | **All patients (N=956)**<br>**n (%)** | **Tested on TAC (N=637)**<br>**n (%)** |
| --- | --- | --- |
| **Gender** |  |  |
| Female | 474 (50) | 319 (50) |
| **Age in years** |  |  |
| Median age (IQR) | 28 (1-44) | 33 (6-47) |
| **Age group in years** |  |  |
| <1 | 232 (24) | 83 (13) |
| 1-4 | 129 (13) | 71 (11) |
| 5-14 | 39 (4) | 24 (4) |
| 15-24 | 45 (5) | 36 (6) |
| 25-44 | 294 (31) | 242 (38) |
| 45-64 | 169 (18) | 143 (22) |
| ≥65 | 48 (5) | 38 (6) |
| **Site province** |  |  |
| Mpumalanga | 335 (35) | 176 (28) |
| North-West | 621 (65) | 461 (72) |
| **Disease presentation** |  |  |
| Acute SRI | 480 (50) | 286 (45) |
| Chronic SRI | 476 (50) | 351 (55) |
| **HIV status** |  |  |
| HIV infected | 468 (49) | 369 (58) |
| HIV uninfected | 478 (50) | 264 (41) |
| Unknown | 10 (1) | 4 (1) |
| **Case fatality** | 51/937 (5) | 42/624 (7) |
| **Symptom duration in days** |  |  |
| Median duration (IQR) | 4 (2-15) | 6 (2-18) |

Chronic SRI defined as ≥10 days symptom duration

### Respiratory pathogens detected using TAC

Among the 637 patients who were tested using TAC, 637 had blood tested, 487 had NP/OP tested and, 411 had sputum tested. At least one pathogen was detected in 83% (527/637) of patients: any bacterial pathogen in 67% (427/637) and any viral pathogen in 47% (302/637). Among all SRI patients, common pathogens detected included *H. influenzae* (225/637; 35%), *S. pneumoniae* (224/637; 35%), rhinovirus (144/637; 23%), *S. aureus* (129/637; 20%), *K. pneumoniae* (85/637; 13%), *M. tuberculosis* (75/637; 12%), and RSV (57/637; 9%) (Tables 2 and S2).

**Table 2:** Bacterial and viral pathogens detected in patients hospitalized with severe respiratory illness by specimen type, at two sentinel sites in South Africa, January – December 2017

| Pathogen | All specimen types<br>(N=637) | NP/OP specimens<br>(N= 487) | Sputum specimens (N=411) | Blood<br>(N=637) |
| --- | --- | --- | --- | --- |
| **Bacteria** |  |  |  |  |
| *H. influenzae* * | 225 (35) | NT | 215 (52) | 23 (4) |
| *S. pneumoniae* * | 224 (35) | NT | 183 (45) | 80 (13) |
| *S. aureus* * | 129 (18) | NT | 104 (25) | 32 (5) |
| *K. pneumoniae* * | 85 (13) | NT | 45 (11) | 45 (7) |
| *M. tuberculosis* | 75 (12) | 18 (4) | 66 (16) | NT |
| Group B streptococcus* | 47 (7) | NT | 35 (9) | 13 (2) |
| *A. baumannii* * | 37 (6) | NT | 19 (5) | 19 (3) |
| *P. aeruginosa* * | 33 (5) | NT | 23 (6) | 12 (2) |
| *Bordetella* spp. | 12 (2) | 3 (1) | 9 (2) | NT |
| *M. pneumoniae* | 11 (2) | 9 (2) | 5 (1) | NT |
| *Legionella* spp. | 8 (1) | 3 (1) | 5 (1) | NT |
| **Viruses** |  |  |  |  |
| Rhinovirus | 144 (23) | 119 (24) | 66 (16) | NT |
| RSV | 57 (9) | 54 (11) | 14 (3) | NT |
| Adenovirus | 47 (7) | 24 (5) | 30 (7) | NT |
| Enterovirus | 37 (6) | 34 (7) | 7 (2) | NT |
| Influenza A | 31 (5) | 25 (5) | 20 (5) | NT |
| hMPV | 20 (3) | 13 (3) | 8 (2) | NT |
| Influenza B | 17 (3) | 5 (1) | 15 (4) | NT |
| Parainfluenza 2 | 12 (2) | 11 (2) | 5 (1) | NT |
| Parainfluenza 1 | 7 (1) | 5 (1) | 4 (1) | NT |
| Parainfluenza 3 | 6 (1) | 3 (1) | 4 (1) | NT |

NP/OP, combined naso-and oro-pharyngeal specimen; hMPV, human metapneumovirus; RSV, respiratory syncytial virus; NT, not tested.
\*For the respiratory TAC, these pathogens were regarded as positive if detected in sputum specimens (N=445).

When comparing pathogen prevalence by age, with the exception of *K. pneumoniae and A. baumannii*, bacterial pathogens were more prevalent in the older children aged ≥5 years and adults compared to the young children aged <5 years (72% [347/483] vs 52% [80/154]; *p* <0.001) (Fig 2) (Table 3). In contrast, *K. pneumoniae* at (19% [30/154] vs 11% [55/483]; *p* =0.01 and *A. baumannii* at 10% [16/154] vs 4% [21/483]; *p* =0.005 were more prevalent in the younger children. The majority of the viral pathogens, with the exception of influenza B, were also detected in the young children (77% [118/154] vs 38% [184/483]; *p* <0.001). Influenza B was only detected in the older children and adults at 4% (17/483).

**Table 3:**
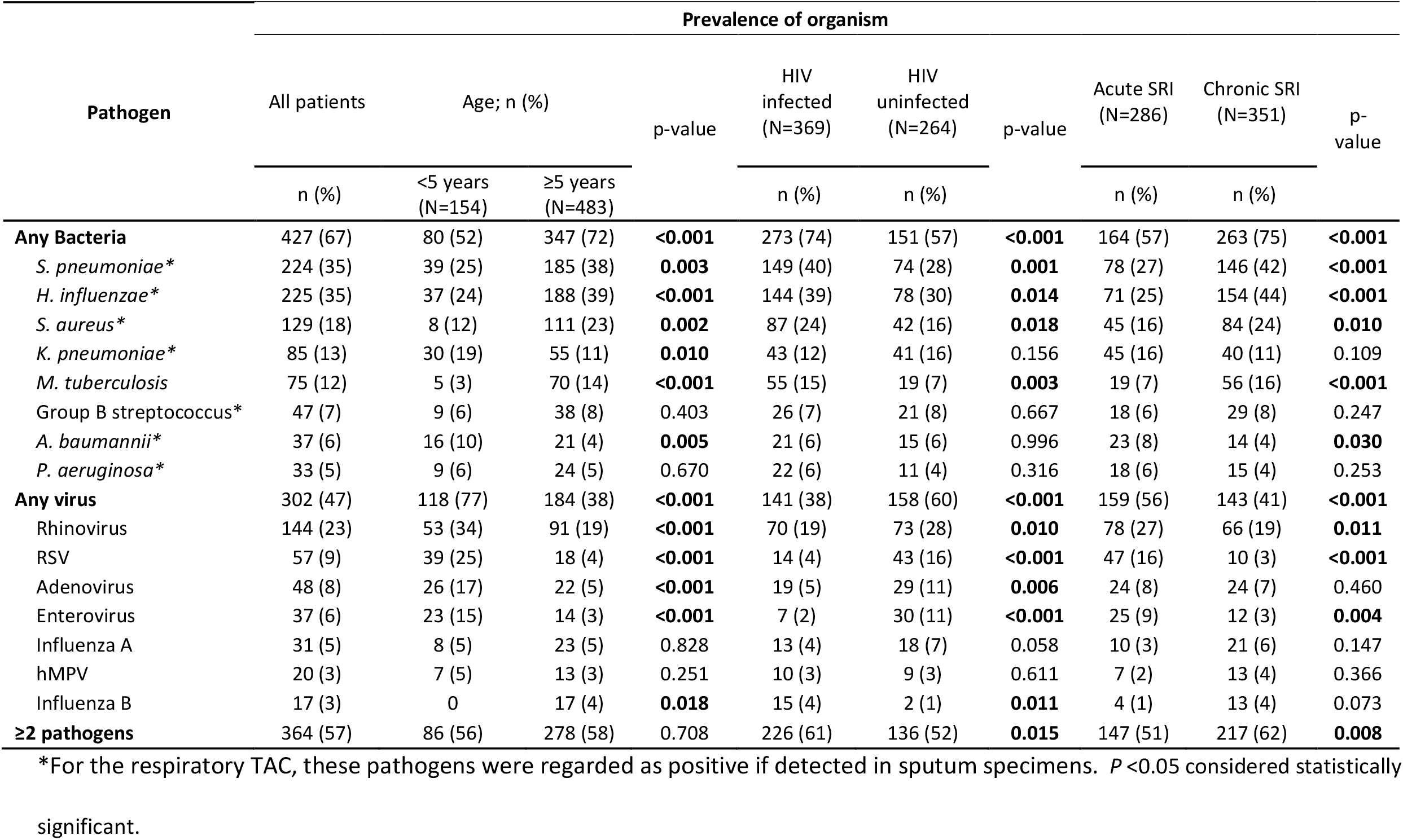
Prevalence of bacterial and viral pathogens, stratified by age, HIV status, and symptom duration, among patients hospitalized with severe respiratory illness, at two sentinel sites in South Africa, January – December 2017

**Fig 2:**
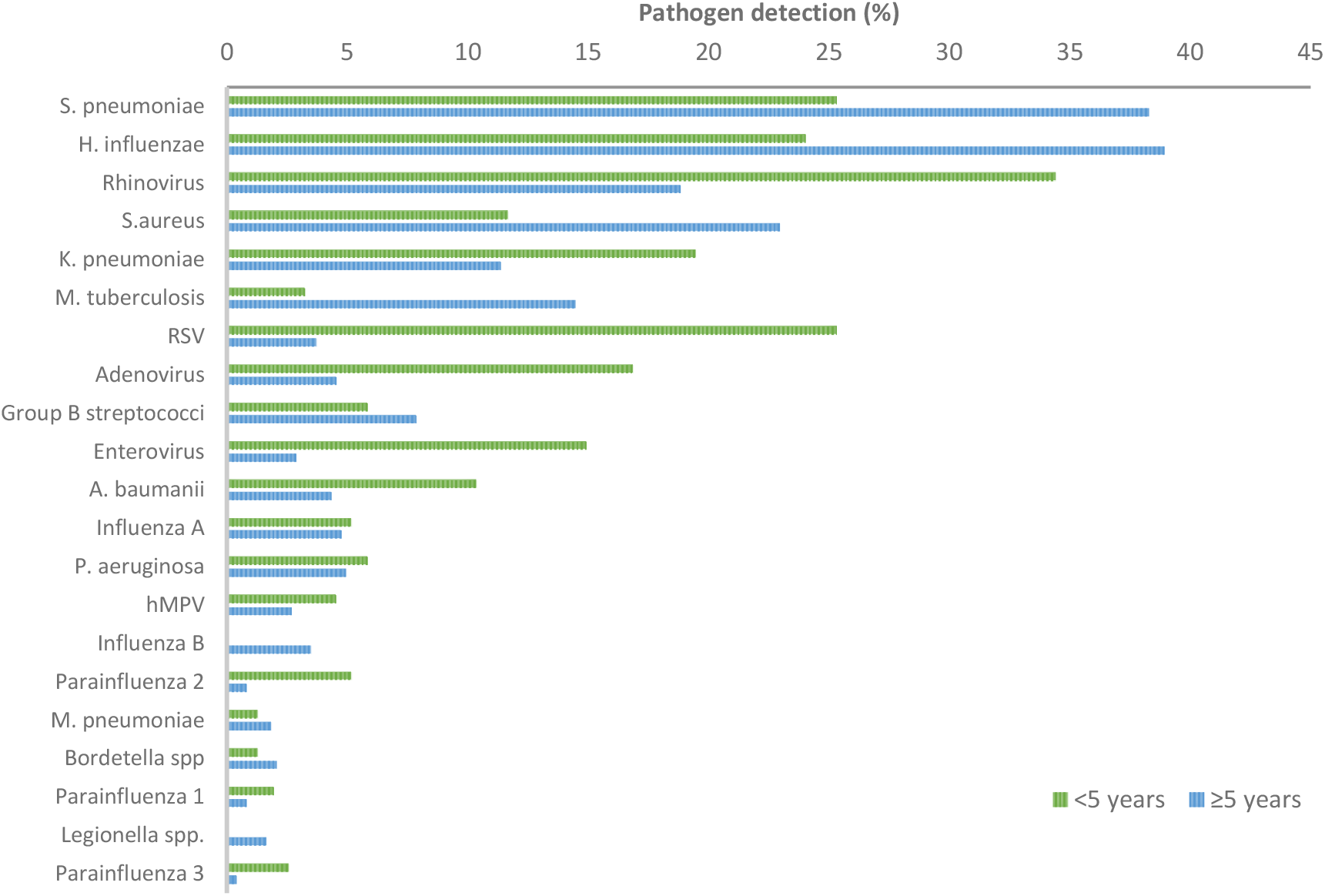
Prevalence of pathogens detected among patients hospitalized with severe respiratory illness using TAC, stratified by age, January – December 2017

Further, when comparing the prevalence of pathogens between HIV-infected and -uninfected patients, it was found that bacterial pathogens were more commonly detected among HIV-infected patients (74% [273/369] vs 57% [151/264]; *p* <0.001) (Table 3). In contrast, the prevalence of viral pathogens was higher among the HIV-uninfected patients (60% [158/264] vs 38% [141/369]; *p* <0.001). Viral pathogens were more common among patients who presented with acute SRI (56% [159/286] vs 41% [143/351]; *p* <0.001) when compared to chronic SRI. In contrast, bacterial pathogens were more common amongst the chronic cases (75% [263/351] vs 57% [164/286]; *p* <0.001) when compared to acute SRI. Among patients with acute SRI, viral pathogens were more common among the younger children (76% [96/127] vs 40% [63/159]; *p* <0.001) compared to older children and adults.

Pathogens were co-detected in 57% (364/637) of the patients, with up to 11 pathogens detected in a single patient. The majority of these were bacterial-viral co-pathogens which were detected in 32% (202/637) of the patients, ≥2 bacterial pathogens in 21% (137/637) and ≥2 viral pathogens in 4% (25/637) (Fig 3). Pathogen co-detection rates were higher among HIV-infected patients (61% [226/369] vs 52% [136/264]; *p* = 0.02) and patients presenting with chronic SRI (62% [217/351] vs 51% [147/286]; *p* = 0.01) (Table 3). A pathogen was detected in 28 (67%) patients among the 42 who died of which 43% (12/28) were bacterial-viral co-pathogens, 39% (11/28) were bacterial and 18% (5/28) were viral pathogens. Pathogen co-detection rates were lower among patients that survived compared to those that died, although not statistically significant (68% [334/492] vs 82% [23/28]; *p* = 0.17).

**Fig 3:**
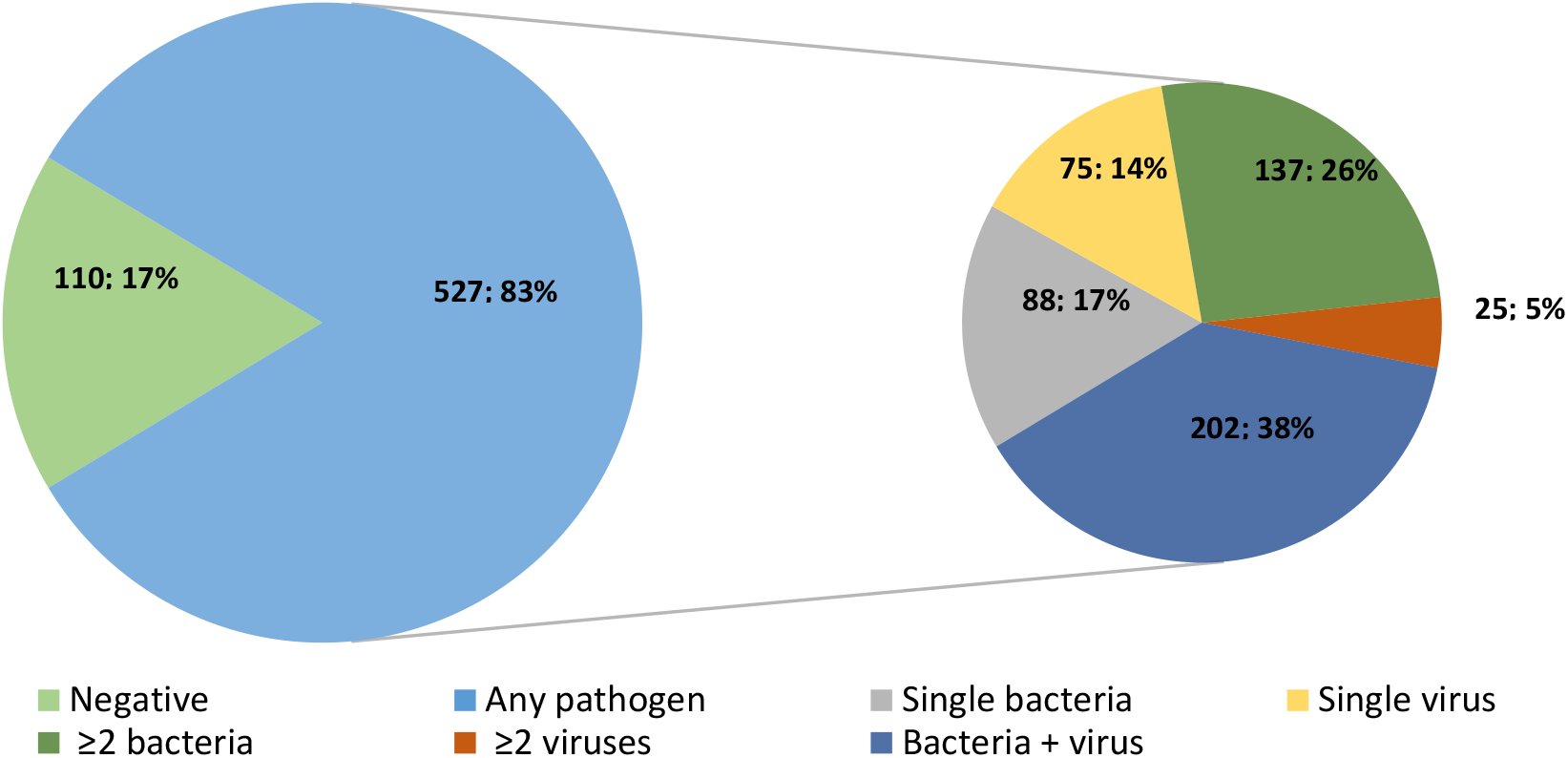
Co-detection of respiratory pathogens among patients hospitalized with severe respiratory illness at two sentinel sites in South Africa, January to December 2017

### Pathogens detected in blood specimens

A bacterial pathogen was detected in 48% (305/637) of blood specimens tested using TAC. *S. pneumoniae* was the most frequently detected pathogen (80/637; 13%) while *K. pneumoniae* (45/637; 7%), *S. aureus* (32/637; 5%), *H. influenzae* (23/637; 4%) and *A. baumannii* (19/637; 3%) were also identified (Table 2). The detection rate of *S. pneumoniae* in young children (17/154; 11%) and older children and adults (63/483; 13%) was comparable (*p* = 0.41). While up to seven bacterial pathogens were detected in a single blood specimen, ≥2 bacterial co-pathogens were only detected in 11% (70/637) of patients (Fig 3).

Both a blood and sputum specimen were tested for 411 patients. Of the patients that tested positive for *H. influenzae* in blood, 93% (13/14) had the same pathogen detected in sputum (Table S3). A similar finding was noted for *S. pneumoniae* (39/54; 72%). In contrast, the rates of positivity between blood and sputum specimens were lower for *S. aureus* (7/20; 30%), *K. pneumoniae* (5/23; 22%) and *A. baumannii* (1/8; 13%). Similar trends were noted in the positivity rates of these pathogens between blood and NP/OP among the 487 patients who had both specimens tested using TAC (Table S3).

### Comparison between TAC and individual real-time PCR (IRTP)

There was moderate agreement (OPA 91%; κ=0.503 [95% CI 0.392-0.614]) in the detection of *S. pneumoniae* between the two assays, in which 34/90 (38%) of the *S. pneumoniae* detected in 634 bloods were detected on both platforms, 44/90 (49%) were detected using TAC only and 12/90 (13%) were detected on individual real-time PCR only (Table 4). There was substantial agreement between the assays in the detection of *Bordetella* spp. in NP/OP (OPA 100%; κ=0.799 [95% CI 0.413-1.000]) and in sputum (OPA 99%; κ=0.618 [95% CI 0.336-0.900]), and almost perfect agreement for the detection of RSV (OPA 97%; κ=0.837 [95% CI 0.757-0.918]), influenza A (OPA 99%; κ=0.857 [95% CI 0.745-0.970]) and influenza B (OPA 100%; κ=0.748 [95% CI 0.411-1.000]) between the two assays. Discrepancies between the assays were noted in specimens with low to medium pathogen load (Ct > 30), in which TAC yielded more pathogens than IRTP (Table S4).

**Table 4:** Comparison of pathogens detected in patients hospitalized with severe respiratory illness using TaqMan Array Card (TAC) and individual real-time PCR

| Pathogen detected | Specimen type | IRTP + TAC+ | IRTP+ TAC- | IRTP- TAC + | Total + | IRTP- TAC- | OPA (%) | $\kappa$ (95%CI) |
| --- | --- | --- | --- | --- | --- | --- | --- | --- |
| *S. pneumoniae* | Blood | 34 | 12 | 44 | 90 | 544 | 91.17 | 0.503 (0.392-0.614) |
| *Bordetella* spp. | NP/OP | 2 | 0 | 1 | 3 | 481 | 99.79 | 0.799 (0.413-1.000) |
|  | Sputum | 5 | 2 | 4 | 11 | 400 | 98.54 | 0.618 (0.336-0.900) |
| RSV | NP/OP | 44 | 5 | 10 | 59 | 428 | 96.92 | 0.837 (0.757-0.918) |
| Influenza A | NP/OP | 19 | 0 | 6 | 25 | 462 | 98.77 | 0.857 (0.745-0.970) |
| Influenza B | NP/OP | 3 | 0 | 2 | 5 | 482 | 99.59 | 0.748 (0.411-1.000) |

NP/OP, combined naso-and oro-pharyngeal specimen, IRTP, individual real-time PCR. RSV,
respiratory syncytial virus. +, positive. -, negative. OPA, overall percent agreement. $\kappa$ , Cohen's
kappa statistic as a measure of agreement: 0.2 as slight agreement, 0.21-0.4 as fair, 0.41-0.60 as
moderate, 0.61-0.80 as substantial, and 0.81-1.00 almost perfect agreement.

## Discussion

We describe the use of a multi-pathogen real-time PCR platform, TAC, for the detection of bacterial and viral pathogens in patients hospitalized with SRI at two sentinel sites in South Africa from January to December 2017. At least one respiratory pathogen was detected in 83% of the patients tested, in which bacterial and viral pathogen detections were 67% and 47%, respectively. Overall, the most commonly detected pathogens included *S. pneumoniae, H. influenzae*, rhinovirus, *S. aureus, K. pneumoniae, M. tuberculosis*, RSV, adenovirus, GBS, *P. aeruginosa*, enterovirus, and *A. baumannii*.

Bacterial pathogens were more commonly detected in patients aged ≥5 years, HIV-infected individuals and patients presenting with chronic symptoms. Among children aged <5 years, *S. pneumoniae* and *H. influenzae* were the two most prevalent bacterial pathogens, with 25% and 24% detection rates, respectively. Similar findings were reported by the Pneumonia Etiology Research for Child Health (PERCH) study in which the detection of *S. pneumoniae* in lower respiratory specimens was 24%-33% while *H. influenzae* (albeit non-type b) was detected at 16% in this age group (12). However, the detection of these pathogens in sputum specimens can also be a result of contamination with upper respiratory tract flora due to poor specimen collection practices. These high detection rates in our study, which were mostly in sputum, may therefore be attributed to high carriage rates of these pathogens in the upper respiratory tract of young children which can range from 7% -93%, and 5% -70%, respectively, in low and middle income countries (27). When restricted to blood specimens only in children <5 years, the detection rate of *S. pneumoniae* was 11% in our study. This 11% prevalence of *S. pneumoniae* in the young children in our study is slightly higher than previously reported in South Africa, in which the prevalence of *S. pneumoniae* was 4%-8% in blood tested using PCR alone or a combination of PCR and culture (3, 12).

Our findings highlight potential roles for *S. aureus, K. pneumoniae, M. tuberculosis*, adenovirus, GBS, enterovirus, *P. aeruginosa, A. baumannii*, and influenza A as important causes of SRI, consistent with previous reports in South Africa, Niger and other African countries between 2010 and 2015 (12, 23, 24, 28, 29). The 12% detection rate of *M. tuberculosis* in our study, the majority of which was detected among older children and adults, is in agreement with a previous report in South Africa of 15% prevalence of the pathogen in patients aged >15 years hospitalized with SRI between 2010 and 2011 (29). We also identified *S. aureus, K. pneumoniae* and *A. baumannii* among the most commonly detected pathogens in agreement with a recent report from the PERCH study as described above (12). In our study, while *K. pneumoniae* and *A. baumannii* were more prevalent in young children, *S. aureus* was more prevalent among older children and adults thus suggesting that these pathogens may play an important role in SRI in these respective age groups.

Viral pathogen detections were more common in young children aged <5 years, HIV-uninfected individuals, and in patients with acute SRI. We detected viruses in 77% of all SRI patients aged <5 years, which is slightly higher than a recent finding by the PERCH study where viral detections accounted for 61% (95% credible interval 57-66) of HIV-uninfected children aged <5 years hospitalized with SRI (12). However, this difference can be attributed to the difference in the analytical approach for inference of SRI etiology as well as study design as in our study, we tested both HIV-infected and -uninfected patients enrolled at only two surveillance sites in South Africa (12). We also noted that the finding of 56% detection of a viral pathogen in our acute SRI patients is lower than previously reported by Pretorius et al. (2016), who reported a 70% prevalence of viruses among patients of all ages hospitalized with acute SRI in South Africa from 2012 to 2015 (30). This is likely due to different age distributions of the study populations (73% were aged <5 years in study by Pretorius et al. compared to 24% in our study) as when restricted to acute SRI patients aged <5 years in our study, viral detection was 76% in this age group.

Rhinovirus and RSV were the most common viral pathogens detected in children aged <5 years consistent with previous reports in the USA, Asia, South Africa and other African countries, ranging from 31%-37% for RSV and 16%-28% for rhinovirus between 2009 and 2015 (12, 24, 31). Rhinovirus was previously reported to have low attributable association (2-fold) with acute SRI and high detection in asymptomatic controls in this age group in South Africa between 2012 and 2015 (30). In our study, rhinovirus was rarely detected as a sole pathogen in this age group (only 25% of all rhinovirus detections in children aged <5 years). Our finding of 25% RSV detection rate in patients aged <5 years with SRI in South Africa is consistent with a previous report by Cohen et al. (2015) who reported a 26% prevalence of RSV in this age group between 2009 and 2012 (3). Pretorius et al. (2016) previously reported a 10-fold odds of detection of RSV in acute SRI cases compared to healthy controls in young children aged <5 years in South Africa between 2012 and 2015 thus highlighting a significant role of this pathogen in childhood SRI (30).

Pathogen co-detections of between two and 11 pathogens in a single patient accounted for 57% of patients tested and were more commonly detected in HIV-infected individuals and patients with chronic SRI. This complicates the ability to assign disease causality warranting further research on the role of mixed infections in severe respiratory disease. Among patients with co-infections, 32% had bacteria-virus, 21% bacteria-bacteria and 4% virus-virus co-detections. Co-detections, especially bacterial-viral, have been reported to be associated with more severe outcomes of SRI (32); in our study however, there was no difference in the rate of bacterial-viral co-detections between the patients who died and those who survived however this may be due to small numbers of patients with a fatal outcome.

TAC and individual real-time PCR assays were compared for the detection of *S. pneumoniae, Bordetella* spp., influenza A and B and RSV A/B. We noted a substantial agreement between the assays for the detection of the pathogens in specimens with higher pathogen load (using Ct value as a proxy for pathogen load). However, in specimens with lower pathogen load, TAC detected more pathogens than individual real-time PCR. As reported previously by Diaz et al. (2013), our findings suggest that the increased number of replicates on TAC may have increased the ability to detect pathogens with low pathogen load (19). However, other methodological differences, such as blood pre-lysis, increased specimen volume, different TNA extraction methods and PCR enzymes likely also contributed to improved TAC pathogen detection.

Overall, use of TAC for the detection of 21 pathogens in a single patient yielded a pathogen in 83% of the patients. Often, etiologic studies for the detection of pneumonia-causing pathogens select, *a priori*, a panel of pathogens that are targeted by currently licensed vaccines or those still in the pipelines as well as pathogens which are traditionally known causes of the disease using conventional methods such as culture. Our findings therefore suggest that use of multi-pathogen panels may be useful to understand the roles of other pathogens in SRI that may have been traditionally underestimated as causes of SRI and which may be of public health importance.

Limitations in our study included that we only considered *S. pneumoniae, S. aureus, K. pneumoniae, H. influenzae, A. baumannii, P. aeruginosa* and GBS to be pathogens if detected in blood and sputum specimens but excluded detection of these pathogens in NP/OP as they are common colonizers of the upper respiratory tract and we could therefore not differentiate between carriage and disease. This exclusion could have inadvertently resulted in the underestimation of these pathogens. Conversely, detection of these pathogens in sputum can also be a result of contamination with upper respiratory tract flora due to poor specimen collection procedures and so may have overestimated the reported detection rates. Also, detection of these pathogens in blood can be as a result of transient bacteremia and thus further overestimating their role in SRI. Lastly, when comparing the performances of TAC and individual real-time PCR for the detection of *S. pneumoniae, Bordetella* spp., influenza A and B and RSV, we did not perform further verification of the discrepant results.

## Conclusion

In our study, the use of a multi-pathogen real-time PCR platform yielded a pathogen in 83% of the SRI patients. It also suggested potentially significant roles for non-vaccine preventable pathogens such as RSV, *S. aureus* and *K. pneumoniae*, in childhood SRI. Surveillance activities often monitor trends in the epidemiology of vaccine-preventable pathogens such as *S. pneumoniae, Bordetella* spp., and influenza, however, our study suggests that monitoring of other pathogens such as *S. aureus* and *K. pneumoniae* should be considered. These data could be useful in informing future preventive or therapeutic public health interventions. Also, as pathogen co-detections were common with the use of multi-pathogen tests, the clinical role of these potential mixed infections in disease requires further investigation.

## Acknowledgements

The authors would like to thank GERMS-SA for the study design, patient enrolment and data collection; the Centre for Respiratory Diseases and Meningitis of the National Institute for Communicable Diseases, a division of the National Health Laboratory Service for laboratory testing and the U.S. CDC for the development and optimisation of the TaqMan Array Card assays.

## Authors’ contributions

MM performed the experiments, interpreted the results, organised the project and drafted the manuscript. CR assisted with laboratory testing. NW supervised the project, interpreted the results, and edited the manuscript. MdP, OH, EV, OM, KN, ST, SW, MW, CC and AvG assisted in the study design and/or patient enrolment, interpreted the results, and edited the manuscript. All authors contributed to manuscript revision, read and approved the submitted version. The authors declare that they have no competing interests; CC has received grant support from Sanofi Pasteur, Advanced Vaccine Initiative, and payment of travel costs from Parexel. NW and AvG have received grant support from Sanofi Pasteur.

