## Supplementary material for "Pathogens detected using a syndromic molecular diagnostic platform in patients hospitalized with severe respiratory illness in South Africa in 2017": Table S1

**Table S3:** Detection of bacteraemia pathogens in paired blood and sputum, and paired blood and NP/OP of patients hospitalized with SRI

| **Pathogen** | **Paired blood and Sputum specimens (N=411)** | | | **Paired blood and NP/OP specimens (N=487)** | | |
|---|---|---|---|---|---|---|
| | **Total Blood +** | **Blood+ Sputum +** | **Blood Only +** | **Total Blood +** | **Blood+ NP/OP +** | **Blood Only +** |
| *S. pneumoniae* | 54 | 39 (72) | 15 | 62 | 46 (74) | 16 |
| *K. pneumoniae* | 23 | 5 (22) | 18 | 38 | 15 (39) | 23 |
| *S. aureus* | 20 | 7 (30) | 13 | 29 | 10 (34) | 19 |
| *H. influenzae* | 14 | 13 (93) | 1 | 20 | 19 (95) | 1 |
| *A. baumannii* | 8 | 1 (13) | 7 | 16 | 4 (25) | 12 |
| Group B streptococci | 4 | 1 (25) | 3 | 13 | 3 (23) | 10 |
| *P. aeruginosa* | 6 | 2 (33) | 4 | 9 | 0 | 9 |

**Table S4:** Comparison of cycle threshold (Ct) values for pathogens detected in patients hospitalized with SRI using TAC and individual real-time PCR (IRTP)

| **Pathogen detected** | **Specimen type** | Ct <30 | | 30≤Ct≤35 | | Ct >35 | | Negative | | Total tested | |
|---|---|---|---|---|---|---|---|---|---|---|---|
| | | **IRTP** | **TAC** | **IRTP** | **TAC** | **IRTP** | **TAC** | **IRTP** | **TAC** | **IRTP** | **TAC** |
| *S. pneumoniae* | B | 5 | 7 | 15 | 30 | 26 | 41 | 588 | 556 | 634 | 634 |
| *Bordetella* spp. | N | 2 | 2 | 0 | 1 | 0 | 0 | 480 | 479 | 482 | 482 |
| | S | 2 | 2 | 0 | 4 | 0 | 3 | 409 | 402 | 411 | 411 |
| RSV | N | 39 | 41 | 9 | 7 | 1 | 6 | 438 | 433 | 487 | 487 |
| Influenza A | N | 17 | 16 | 2 | 3 | 0 | 6 | 468 | 462 | 487 | 487 |
| Influenza B | N | 2 | 2 | 2 | 3 | 0 | 0 | 483 | 482 | 487 | 487 |

Ct <30, high pathogen load. Ct 30-35, medium pathogen load. Ct > 35, low pathogen load. B, blood, N, combined naso-and oro-pharyngeal specimen in universal transport media, S, sputum. IRTP, individual real-time PCR. RSV, respiratory syncytial virus.
