## Supplementary material for "Pathogens detected using a syndromic molecular diagnostic platform in patients hospitalized with severe respiratory illness in South Africa in 2017": Table S2

**Table S2:** Pathogens detected in respiratory specimens of patients hospitalized with SRI using TaqMan Array Card (TAC)

| Pathogen detected | Specimen type | **All patients** | | | | Patients aged <5 years | | | | Patients aged ≥5 years | | | |
|---|---|---|---|---|---|---|---|---|---|---|---|---|---|
| | | **TAC+** | **TAC-** | **Total** | **Pathogen detection (%)** | **TAC+** | **TAC-** | **Total** | **Pathogen detection (%)** | **TAC+** | **TAC-** | **Total** | **Pathogen detection (%)** |
| S. pneumoniae | B+S | 224 | 413 | 637 | 35 | 39 | 115 | 154 | 25 | 185 | 298 | 483 | 38 |
| *H. influenzae* | B+S | 225 | 412 | 637 | 35 | 37 | 117 | 154 | 24 | 188 | 295 | 483 | 39 |
| Rhinovirus | N+S | 144 | 493 | 637 | 23 | 53 | 101 | 154 | 34 | 91 | 392 | 483 | 19 |
| *S.aureus* | B+S | 129 | 508 | 637 | 20 | 18 | 136 | 154 | 12 | 111 | 372 | 483 | 23 |
| *K. pneumoniae* | B+S | 85 | 552 | 637 | 13 | 30 | 124 | 154 | 19 | 55 | 428 | 483 | 11 |
| *M. tuberculosis* | N+S | 75 | 562 | 637 | 12 | 5 | 149 | 154 | 3 | 70 | 413 | 483 | 14 |
| RSV | N+S | 57 | 580 | 637 | 9 | 39 | 115 | 154 | 25 | 18 | 465 | 483 | 4 |
| Adenovirus | N+S | 48 | 589 | 637 | 8 | 26 | 128 | 154 | 17 | 22 | 461 | 483 | 5 |
| Group B streptococci | B+S | 47 | 590 | 637 | 7 | 9 | 145 | 154 | 6 | 38 | 445 | 483 | 8 |
| Enterovirus | N+S | 37 | 600 | 637 | 6 | 23 | 131 | 154 | 15 | 14 | 469 | 483 | 3 |
| *A. baumanii* | B+S | 37 | 600 | 637 | 6 | 16 | 138 | 154 | 10 | 21 | 462 | 483 | 4 |
| Influenza A | N+S | 31 | 606 | 637 | 5 | 8 | 146 | 154 | 5 | 23 | 460 | 483 | 5 |
| *P. aeruginosa* | B+S | 33 | 604 | 637 | 5 | 9 | 145 | 154 | 6 | 24 | 459 | 483 | 5 |
| HMPV | N+S | 20 | 617 | 637 | 3 | 7 | 147 | 154 | 5 | 13 | 470 | 483 | 3 |
| Influenza B | N+S | 17 | 620 | 637 | 3 | 0 | 154 | 154 | 0 | 17 | 466 | 483 | 4 |
| Parainfluenza 2 | N+S | 12 | 625 | 637 | 2 | 8 | 146 | 154 | 5 | 4 | 479 | 483 | 1 |
| *M. pneumoniae* | N+S | 11 | 626 | 637 | 2 | 2 | 152 | 154 | 1 | 9 | 474 | 483 | 2 |
| Bordetella spp | N+S | 12 | 625 | 637 | 2 | 2 | 152 | 154 | 1 | 10 | 473 | 483 | 2 |
| Parainfluenza 1 | N+S | 7 | 630 | 637 | 1 | 3 | 151 | 154 | 2 | 4 | 479 | 483 | 1 |
| *Legionella* spp. | N+S | 8 | 629 | 637 | 1 | 0 | 154 | 154 | 0 | 8 | 475 | 483 | 2 |
| Parainfluenza 3 | N+S | 6 | 631 | 637 | 1 | 4 | 150 | 154 | 3 | 2 | 481 | 483 | 0 |

Specimen types N, naso- /oropharyngeal specimens (NP/OP); S, sputum.
