## Supplementary material for "Pathogens detected using a syndromic molecular diagnostic platform in patients hospitalized with severe respiratory illness in South Africa in 2017": Table S3 and Table S4

**Table S1:** List of organisms tested by Taqman Array Card (TAC) in blood and respiratory specimens

| **Organism** | **Respiratory TAC**[a] | **Blood TAC**[a] |
|---|---|---|
| *Acinetobacter baumannii* | ✓ | ✓ |
| *Bordetella pertussis* | ✓ | |
| *Escherichia coli / Shigella* spp. | ✓ | ✓ |
| *Group B Streptococcus* | ✓ | ✓ |
| *Haemophilus influenzae* | ✓ | ✓ |
| *Klebsiella penumoniae* | ✓ | ✓ |
| *Legionella* spp. | ✓ | |
| *Mycobacterium tuberculosis* | ✓ | |
| *Mycoplasma pneumoniae* | ✓ | |
| *Pseudomonas aeruginosa* | ✓ | ✓ |
| *Staphylococcus aureus* | ✓ | ✓ |
| *Streptococcus pneumoniae* | ✓ | ✓ |
| Adenovirus | ✓ | |
| Cytomegalovirus | | ✓ |
| Enterovirus | ✓ | |
| Human metapneumovirus | ✓ | |
| Influenza A | ✓ | |
| Influenza B | ✓ | |
| Parainfluenza virus 1 | ✓ | |
| Parainfluenza virus 2 | ✓ | |
| Parainfluenza virus 3 | ✓ | |
| Respiratory syncytial virus | ✓ | |
| Rhinovirus | ✓ | |

[a] All targets were present in two replicates on the respiratory TAC or four replicates on the blood TAC, with the exception of *H. influenzae* which was present in two replicates on the blood TAC.
